# Human-AI Collaboration in Clinical Reasoning: A UK Replication and Interaction Analysis

**DOI:** 10.1101/2025.08.25.25334383

**Authors:** J Healy, J Kossoff, M Lee, C Hasford

## Abstract

**Objective:** A paper from Goh et al found that a large language model (LLM) working alone outperformed American clinicians assisted by the same LLM in diagnostic reasoning tests [1]. We aimed to replicate this result in a UK setting and explore how interactions with the LLM might explain the observed gaps in performance.

**Methods and Analysis:** This was a within-subjects study of UK physicians. 22 participants answered structured questions on 4 clinical vignettes. For 2 cases physicians had access to an LLM via a custom-built web-application. Results were analysed using a mixed-effects model accounting for case difficulty and the variability of clinicians at baseline. Qualitative analysis involved coding of participant-LLM interaction logs and evaluating the rates of LLM use per question.

**Results:** Physicians with LLM assistance scored significantly lower than the LLM alone (mean difference 21.3 percentage points, p < 0.001). Access to the LLM was associated with improved physician performance compared to using conventional resources (73.7% vs 66.3%, p = 0.001). There was significant heterogeneity in the degree of LLM-assisted improvement (SD 10.4%). Qualitative analysis revealed that only 30% of case questions were directly posed to the LLM, which suggests that under-utilisation of the LLM contributed to the observed performance gap.

**Conclusion:** While access to an LLM can improve diagnostic accuracy, realising the full potential of human-AI collaboration may require a focus on training clinicians to integrate these tools into their cognitive workflows and on designing systems that make these integrations the default rather than an optional extra.

## Introduction

Large language models (LLMs) have shown impressive performance across various medical settings and use cases, leading to an explosion of research interest. Initial strong performance on medical exams [3] has prompted investigation into their abilities in a broad range of medical tasks, with triage [4] physician queries [5] and direct patient interactions [6] all promising areas. Diagnostic reasoning represents a fundamental component of clinical practice, and research here is advancing rapidly [7,8].

The current generally accepted paradigm for LLM-integration in clinical practice involves the models supporting human diagnosticians. Goh et al [1] published a surprising result in this regard in 2024. In a study testing diagnostic reasoning they demonstrated that an LLM alone outperformed American physicians using an LLM, and that the use of an LLM did not improve physician performance compared to using conventional resources.

These results are unexpected. Doctors using the LLM have, by definition, access to the answers provided by the LLM and so it is not clear why adding a human clinician would degrade performance. It has been a hypothesis of informatics research dating back to 1960 that the ‘man-computer symbiosis’ will enable individuals to “think as no human brain has ever thought and process data in a way not approached by the information-handling machines” [9]. The fact that the human-LLM interface seems to violate this principle demands further research, but this is an area that has thus far been minimally studied.

It is important to see if this finding extends to other medical contexts. LLMs, as currently designed, are trained on a single corpus of knowledge and are yet deployed across a variety of different clinical contexts and medical cultures. Furthermore, it is important to understand why the addition of humans seems to degrade AI performance. This understanding can inform the design of safer and more effective human-AI integration strategies.

The primary objective was to replicate the finding that a large language model, acting independently, outperforms physicians who have discretionary access to the same model while solving challenging clinical diagnostic reasoning problems. Secondary exploratory qualitative analyses explored how variability in physicians’ prompting choices might mediate any observed performance gap.

### Ethical Approval

The study authors discussed the proposed protocol with the local ethics committee of University College Hospital who agree that the nature of the study (no randomisation, no patient involvement) meant that formal ethics approval was not needed. Informed consent was obtained from all participants and all data was stored and managed in accordance with General Data Protection Regulation (GDPR).

## Methods

### Study Design

This is a within-subjects quasi-experimental trial with an external LLM-only benchmark. Free-text data was collected through a custom-built web application which connected participants to GPT-4o and allowed researchers to retrospectively analyse free-text interactions.

### Recruitment

Twenty-two physicians with at least 1 year of clinical experience were recruited via local mailing lists and posters around UCLH. To compensate them for their time each participant was paid £10 and they were entered into a £200 prize draw, which was allocated randomly at the end of the study. Study recruitment and data collection took place from April 24 2025 to June 11 2025.

### Clinical Vignettes

The same clinical vignettes used by Goh 2024 were selected by the study authors for inclusion in this study. These vignettes and the scoring rubrics have been previously validated for testing clinical reasoning in other contexts [1, 2]. Differences in lab values or medical terminology were converted into values or language in keeping with British medical practice.

To control for the effects of case difficulty and content, the order in which the four clinical vignettes were presented to each participant was randomized. The use of either conventional resources or LLM assistance was administered in a fixed, alternating sequence. Conventional resources included any non-LLM resource available to clinicians, either on-line or off-line (for example, textbooks, BMJ learning, UpToDate, etc). For each participant the first and third cases they encountered were completed using conventional resources, while the second and fourth cases participants had the option of using the study LLM for assistance.

### Data Collection

The study was conducted using a survey tool (Qualtrics). Participants were instructed to access the LLM via a custom-built web application and were aware that qualitative data on their LLM usage was being collected.

The web application was a custom-built tool that called an Application Programming Interface (API) to OpenAI’s GPT-4o model. This model was chosen as the original Goh 2024 paper also used an OpenAI model. No specific prompts were given to the model used by participants and it was not preloaded with any information about the cases or the study more generally.

Study participation was performed asynchronously, with participants briefed both directly and with a pre study information pack.

### Scoring

The mark scheme was replicated from Goh 2024, with a maximum score of 19 for each case (Appendix). A gold standard mark scheme was developed in advance for each case. The cases were then marked using the scheme by two authors (JK, ML). A third author (JH) adjudicated any discrepancy in marks awarded. Inter-rater reliability (prior to adjudication) was assessed using a two-way random-effects, absolute-agreement intraclass correlation for the average of two raters ICC(2,2). Reliability for the averaged score was 0.908, 95% CI [0.870, 0.940], indicating excellent agreement.

The LLM benchmark was derived by prompting GPT-4o to respond to the same cases and questions posed to human clinicians (Appendix for LLM prompt). Markers were blinded as to whether the participants had access to LLMs for specific cases and they were also blinded as to which study IDs represented entirely LLM-generated content.

### Study Outcomes

The primary outcome was the final score as a percentage across all components of the structured clinical reasoning assessment. We aimed to compare LLM-assisted performance with theLLM-only benchmark. Secondary outcomes included the impact of LLMs on clinician performance and a qualitative analysis of the different LLM interaction techniques used by participants.

### Sample-size rationale

A formal power analysis was conducted to determine the sample size required to detect a clinically significant difference between the performance of an LLM-assisted group and an LLM only benchmark. In Goh 2024 this difference was found to be 16%. Assuming a one-sample test at a significance level of 0.05, our sample of 44 cases, clustered within 22 clinicians, provides over 85% power to detect a 16% absolute difference in performance. This calculation accounts for the potential clustering effect by assuming a conservative intraclass correlation coefficient (ICC) of 0.5.

Given the exploratory nature of this work, formal sample size calculations were not undertaken for the qualitative analysis.

### Qualitative Analysis

An a priori coding frame comprising six interaction types was developed prior to analysis (Appendix); these interaction types covered the information that was given to the LLM, as well as the type of questions asked to the LLM, we also captured if participants prompted the model with a specific persona. One author applied the frame to all logs and a second author double-coded 10 % of transcripts to ensure reliability and consistency.

These codes were then tested to see if specific patterns of use were associated with LLM-assisted performance. The nature of this analysis, and the relatively small sample size of participants, means that these findings are strictly exploratory.

We also conducted a detailed analysis of overall LLM use amongst participants. This was done by confirming if the LLM was used at all for the individual case and then determining if the users asked the LLM the specific questions posed by the case.

### Statistical Analysis

To evaluate how closely participants approached the performance of the LLM alone, we compared each participant’s score on the two cases they completed with LLM support to the LLM-only average for those same cases. We then tested whether this was different from zero using a one-sample t-test.

To compare diagnostic performance within-participants with and without LLM-assistance we analyzed case-level scores using a linear mixed-effects model. This model had ‘participant’ as a random intercept and fixed effects for LLM access (Assisted vs Unassisted) and case (four levels). As LLM access was deterministically assigned by position, we did not include position in the model. We report the mean difference (Assisted - Unassisted) with 95% confidence intervals and p-values (Satterthwaite). As the order of LLM-assistance was not randomised, the model outputs should be interpreted cautiously, to further probe the potential effects of order we conducted sensitivity analyses dropping position 2 or 4 and performed leave-one-case-out analyses.

For the qualitative analysis, we recorded the presence or absence of each interaction type and examined associations with performance scores. We dichotomised the volume of interactions into high and low based on whether the number interactions were above or below the median. We employed Welch’s two-sample t-test to test whether these qualitative factors were associated with significant differences in performance. Given there were multiple comparisons being made across the same database a False Discovery Rate (FDR) correction was applied.

All statistical analysis was conducted in R.

## Results

22 clinicians were recruited in total. Of these 21 completed all 4 cases. One clinician completed only 3 cases (1 LLM-assisted, 2 non-LLM assisted), their results are included in the analysis below. Even accounting for this missing case, the study retained a power of over 83% to detect a difference between LLM-assisted and LLM-alone scores.

Table 1 shows the demographics of the study participants. All participants had completed at least one year of post-graduate training. These demographic details are included for background information only, sub-group analysis was not performed for different group performance as the study was not adequately powered for this level of analysis.

**Table 1.** Participant characteristics.

| **Table 1. Participant characteristics** |  |
| --- | --- |
| **Characteristic** | **Number (%)** |
| **Grade** |  |
| Foundation Year 2 - Core Trainee or equivalent (typically 2-4 years clinical experience) | 14 (63.6) |
| Specialist Trainee 3+ (typically 5+ years post-graduate clinical experience) | 7 (31.8) |
| Consultant (completed speciality medical training) | 1 (4.5) |
| **LLM Use** |  |
| I use LLMs frequently (weekly or more) | 6 (27.3) |
| I use LLMs occasionally (more than once a month, less than weekly) | 5 (22.7) |
| I use LLMs rarely (less than once a month) | 6 (27.3) |
| I've never used an LLM | 5 (22.7) |

**Table 2.** Difference between LLM-only and Human LLM-assisted results.

| **Table 2. Difference between LLM-only and Human LLM-assisted results** |  |  |
| --- | --- | --- |
| **Total cases** | **Mean difference [95% CI]** | **p-value** |
| N = 43 | 21.3 percentage points (pp) [13.1pp - 29.5pp] | <0.001 |

**Table 3.** Performance outcomes with and without LLM, mixed-effects model outputs.

|  |  |  |  |
| --- | --- | --- | --- |
| **Group** | **Physician +**<br>**Conventional Resources**<br>**Mean [95% CI]** | **Physician + LLM**<br>**Mean [95% CI]** | **P-value** |
| All participants | 65.3%<br>[58.2% - 72.4%] | 74.4%<br>[67.3% - 81.5%] | 0.001 |

**Table 4.** Usage of the LLM amongst total cases (n = 43)

| **Table 4. Usage of the LLM amongst total cases (n = 43)** |  |
| --- | --- |
| **Use** | **Number (%)** |
| LLM used | 39 (90.7) |
| LLM not used | 4 (9.3) |

### Quantitative Results

Three LLM-only series of responses were submitted for scoring amongst the human-clinician responses. The LLM-only reference scores were: Case 1 = 94.74%, Case 2 = 98.25%, Case 3 = 100.00%, Case 4 = 89.47%. Across all participants, the LLM significantly outperformed human participants by 21.25 percentage points (pp) (95% CI 13.15-29.5pp, p <0.001).

Across all 22 participants, the raw mean score on cases completed with LLM assistance was 73.7%, compared to 66.3% on cases completed with conventional resources. We fit a linear mixed-effect model including mixed effects for case, with a random intercept for participants. With this model, the adjusted score without LLM-assistance was 65.3% (95% CI 58.2-72.4), and with LLM-assistance 74.4% (95% CI 67.3-81.5). The case-adjusted difference was +9.0 percentage points (pp) (95% CI 3.6-14.6pp, p = 0.001). Sensitivity analyses of the impact of case ordering, through dropping specific cases or positions, were consistent with this finding (Appendix). As mentioned in the methodology, the strength of this conclusion is tempered by the fact that the access to the LLM was of a fixed order throughout all participants.

There was a strong positive correlation between LLM-assisted and conventional resource test scores (r = 0.79, 95% CI: 0.55-0.91, p < 0.001; Figure 1). This indicates that participants who performed well using conventional resources also tended to perform well when using LLM assistance, and vice versa.

**Figure 1.**
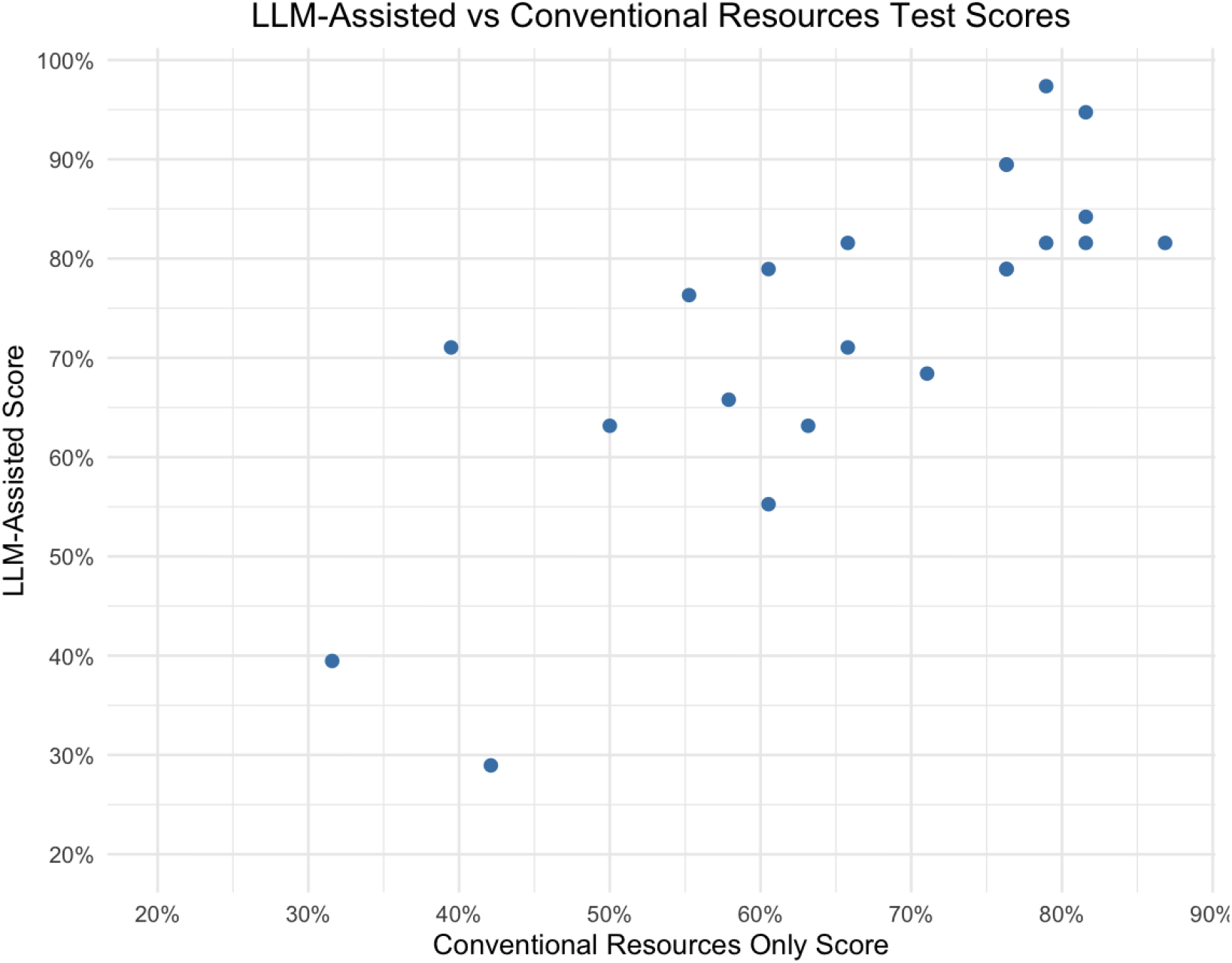
Graph Comparing LLM-Assisted with Conventional Resources Scores

Individual improvements in diagnostic accuracy with LLM assistance demonstrated considerable variability across participants. The mean improvement was 7.42%. Individual performance changes ranged from −13.16% to +31.58%, with a standard deviation of 10.39%, indicating substantial heterogeneity in individual responses to LLM assistance. Figure 2 illustrates that while most clinicians saw improvements in their performance with LLMs, several individuals demonstrated unexpected decreases in performance and the improvements varied significantly between individuals.

**Figure 2.**
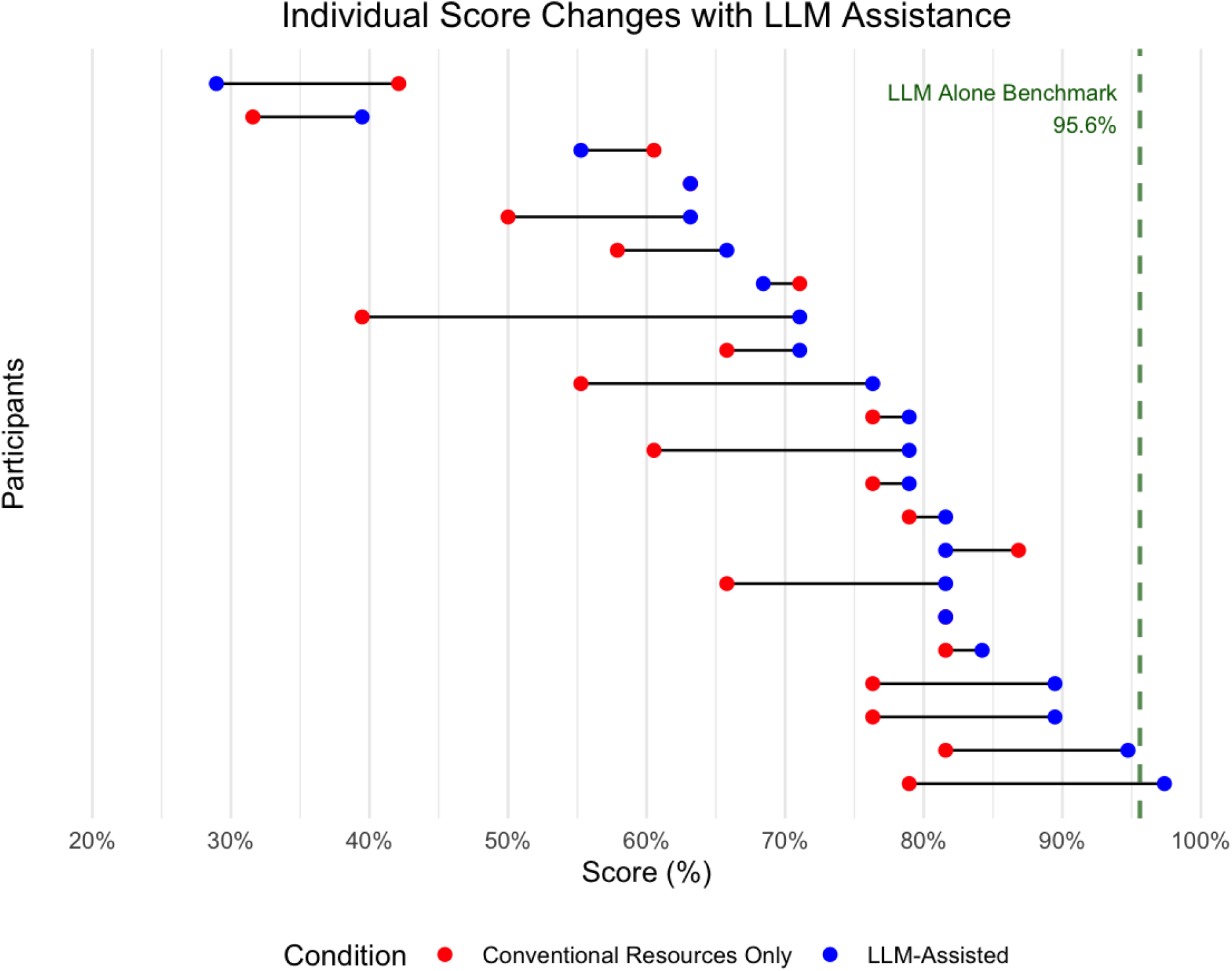
Change in score change with and without LLM

### Qualitative analysis

As an initial screening of LLM use amongst participants, we examined what proportion of cases had any LLM input; defined as a participant asking the LLM a question relating to the case (Table 5). Every participant used the LLM for at least one case, although four participants used the LLM for one case but not for the other (Table 5).

**Table 5.** Qualitative codes impact on overall LLM assisted score.

| **Table 5. Qualitative codes impact on overall LLM assisted score** |  |  |  |  |  |
| --- | --- | --- | --- | --- | --- |
| **Code** | **Code Description** | **Mean Score**<br>**With Code (%)** | **Mean Score**<br>**Without Code (%)** | **p-value** | **p-value**<br>**(FDR)** |
| 1 | Verbatim case content used | 69.41 | 76.13 | 0.412 | 0.950 |
| 2 | Summarised case content used | 77.94 | 67.55 | 0.205 | 0.950 |
| 3 | Verbatim case question used at least once | 71.05 | 74.10 | 0.716 | 0.950 |
| 4 | Exploratory questioning used at least once | 81.29 | 68.42 | 0.047* | 0.568 |
| 5 | Factual questioning used at least once | 76.56 | 70.82 | 0.431 | 0.950 |
| Message volume | High volume (n > 6 messages) | 71.29 | 76.08 | 0.513 | 0.950 |

While every participant used the LLM at some point, only one participant explicitly used the LLM for each question posed in the case (Figure 3). Across all participants only 30% of the questions were posed to the LLM directly, and 27% of participants didn’t pose any of the questions directly to the LLM. The relationship between LLM use and final score was weakly positive (r = 0.26, 95% CI: –0.19 to 0.61, p = 0.25), while this was not statistically significant the sample size for this was small and is heavily confounded by other factors (such as baseline physician performance).

**Figure 3.**
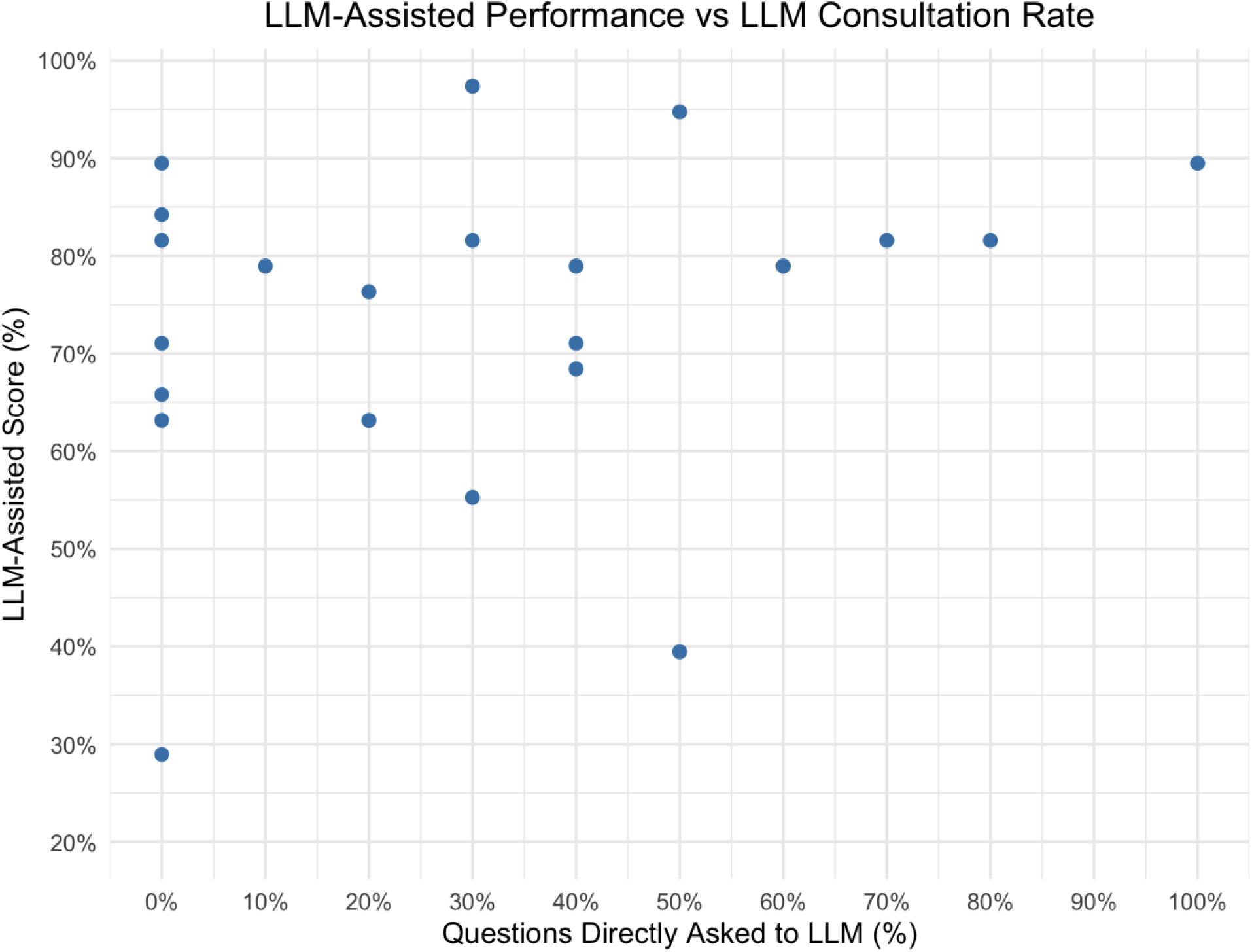
Figure comparing LLM-assisted performance with the proportion of case questions directly posed to the LLM by human participants.

These results are in keeping with the hypothesis that the ‘LLM-assisted’ performance was broadly assessing baseline human clinician performance with only a small incremental benefit from the LLM.

For those occasions where the LLM was used we identified significant variation in strategies applied to the LLMs, characterised by the large variance in the number of messages sent per participant which ranged from 2 to 22 (median = 5.5, IQR = 4.0-9.0). For the purposes of statistical analysis the volume of messages sent was dichotomised into high/low volume, with 6 or more messages sent being characterised as ‘high volume’ and 5 or fewer representing ‘low volume’.

Participants utilised the LLM in a variety of different ways; from simply copying the verbatim case and associated questions, through to exploring specific concepts in a human-LLM dialogue. To capture this variation all participant-LLM interactions were coded using the predefined coding rubric (Appendix). A random 10% of the codes were cross-validated by a second reviewer who found 100% concordance with the original coding, Cohen’s κ = 1.00.

In the initial analysis physicians who asked conceptual or exploratory questions scored significantly higher than physicians who did not (81.3% v 68.4%, p = 0.047, see Table 6). Although the significance of all codes (including conceptual prompting) disappeared with a False Discovery Rate correction for multiple hypotheses, this exploratory piece of analysis suggests that a more in-depth engagement with the material produces a higher quality output from the LLM.

***Code 6 not tested, only used by one participant***.

## Discussion

In this UK replication of Goh 2024’s diagnostic experiment [1], we found once again that a large language model acting alone significantly outperforms clinicians with discretionary access to the same model. We extend Goh’s analysis by finding that an important contributory factor was that participants simply did not use the model when it was available; LLM-assisted performance was heavily correlated with non-LLM-assisted performance and very few participants explicitly asked the LLM for its input on each element of cases.

The most obvious explanation of this under-utilisation is that participants may not have felt the need to involve the LLM; they were likely unaware that the LLM assistance would provide such a significant boost to their performance and considered their initial answers as being sufficient. It is possible that the design of the study (participants were directed to an external custom-built website to use the LLM) may have deterred consistent use, however the fact that the LLM was used in some capacity in almost every case suggests that accessibility was not a significant factor in determining utilisation.

Exploring the prompting strategies in more detail found that participants who used ‘conceptual exploration’ seemed to score higher than participants who did not. While the significance of this relationship disappears with a statistical correction for multiple comparisons, more dedicated research into the clinician-AI interface should explore this potential relationship in more depth.

In contrast to Goh 2024, we found that access to an LLM was associated with significantly improved human performance in diagnostic reasoning cases however this was not a homogenous improvement and there was wide variability in the impact of LLMs on clinician performance. This difference is perhaps explained by the fact that our participants had access to a stronger model (GPT-4o rather than GPT 4) and the fact that there is perhaps a greater understanding of the value of LLMs than there was at the time of Goh 2024 which was originally conducted in 2023. The strength of this conclusion is tempered by the fact that the order of LLM-assistance to non-assisted were fixed in the study design, our efforts to mitigate this through a mixed-effects model and extensive sensitivity analysis cannot fully account for this potential confounding effect.

This study had multiple limitations, which limits generalisability to other settings. The sample size was small and skewed towards clinicians in the earlier stages of their postgraduate training. To improve participant recruitment, the study was conducted asynchronously without direct observation and this means that we could not control for factors such as time spent on each case. Furthermore the marking rubric, while validated in the pre-LLM era, is naturally biased to LLMs which are adept at generating multiple ideas when asked. Finally, the use of clinical vignettes in this lab-based testing environment does not reflect the real-world cognitive work done by clinicians.

This study reinforces that the path to effective AI integration in clinical practice requires an increasing focus on the human user. There is a growing body of work that explores the ‘human in the loop’ model of clinician-AI interactions [10,11] and this work emphasises that future frameworks requiring a ‘human in the loop’ will need to be carefully designed so that these humans are able to best make use of these powerful models. Future work should focus not just on how these systems can be designed, but also on the qualitative aspects of the human-AI interface. In the shorter term, this work suggests that there will be an increasing need to better educate physicians on using AI effectively, in particular educating them on how to prompt effectively, critically appraise LLM outputs, and overcome the cognitive biases that currently prevent them from making the best use of these tools.

### Declaration of generative AI and AI-assisted technologies in the writing process

During the preparation of this work the author(s) used OpenAI’s ChatGPT-5 (Thinking Model) in order to provide feedback on an earlier draft. After using this tool, the authors reviewed and edited the content as needed and take full responsibility for the content of the publication.

## Data Availability

All data produced in the present study are available upon reasonable request to the authors

## Acknowledgements

*We would like to thank Dr Ethan Goh and Dr Adam Rodman for their support in sharing resources and guidance. Without their contributions this replication paper would not have been possible*.

